# Rehabilitation with and without Robot and Allied Digital Technologies (RADTs) in stroke patients: a study protocol for a multicentre Randomised Controlled Trial on the effectiveness, acceptability, usability, and economic-organizational sustainability of RADTs from subacute to chronic phase (STROKEFIT4)

**DOI:** 10.1101/2024.09.11.24313413

**Authors:** Irene Giovanna Aprile, Marco Germanotta, Alessio Fasano, Mariacristina Siotto, Maria Cristina Mauro, Arianna Pavan, Giovanna Nicora, Giuseppina Sgandurra, Alberto Malovini, Letizia Oreni, Nevio Dubbini, Enea Parimbelli, Giovanni Comandè, Christian Lunetta, Pietro Fiore, Roberto De Icco, Carlo Trompetto, Leopoldo Trieste, Giuseppe Turchetti, Silvana Quaglini, the STROKEFIT4 study group

**Affiliations:** IRCCS Fondazione Don Carlo Gnocchi ONLUS, Florence, Italy; Department of Electrical, Computer and Biomedical Engineering, University of Pavia, 27100 Pavia, Italy; Developmental Neurology and Neurorehabilitation Unit, IRCCS Stella Maris Foundation, Pisa, Italy; Department of Clinical and Experimental Medicine, University of Pisa, Pisa, Italy; Laboratory of Medical Informatics and Artificial Intelligence of the Institute of Pavia, Istituti Clinici Scientifici Maugeri IRCCS, Pavia, Italy; IRCCS Fondazione Don Carlo Gnocchi ONLUS, Milan, Italy; Miningful srls, 56121 Pisa, Italy; Sant’Anna School of Advanced Studies, 56127 Pisa Italy; Istituti Clinici Scientifici Maugeri IRCCS, Neurorehabilitation Unit of Milan Institute, Milan, Italy; Istituti Clinici Scientifici Maugeri IRCCS, Neurorehabilitation Unit of Bari Institute, Bari, Italy; Department of Physical and Rehabilitation Medicine, University of Foggia, 71122 Foggia, Italy; Department of Brain and Behavioral Sciences, University of Pavia, Pavia, Italy; Movement Analysis Research Section, IRCCS Mondino Foundation, Pavia, Italy; Department of Neuroscience, Rehabilitation, Ophthalmology, Genetics, Maternal and Child Health, University of Genoa, Italy; IRCCS Ospedale Policlinico San Martino, Genova, Italy; Institute of Management, Scuola Superiore Sant’Anna, 56127 Pisa, Italy

**Keywords:** stroke, rehabilitation, robotics, pragmatic clinical trial, cost-effectiveness analysis

## Abstract

**Introduction:** Rehabilitation after stroke often employs Robots and Allied Digital Technologies (RADTs). However, evidence of their effectiveness remains inconclusive due to study heterogeneity and limited sample sizes. Here, we describe a protocol for a multicentre randomized controlled pragmatic trial aimed at comprehensively and accurately assessing the effectiveness and sustainability of RADT-mediated rehabilitation, compared to traditional rehabilitation.

**Methods and analysis:** This is a pragmatic multicentre, multimodal, randomised, controlled, parallel-group (1:1) interventional study with blinded assessors. The trial will recruit 596 adult post-stroke patients in the subacute phase (less than 6 months post-stroke). Patients will be recruited from thirteen rehabilitation centres participating in a national research initiative, encompassing both outpatient and inpatient clinical settings. Participants will be randomized into either the experimental group, or the control group. The experimental group will receive rehabilitation using RADTs within a new organizational model, where two physical therapists supervise four to six patients; patients will undergo a comprehensive rehabilitation treatment, targeting the following domains: a) upper limb sensorimotor abilities; b) lower limb sensorimotor abilities and gait; c) balance; d) cognitive abilities. In the control group, patients will undergo individual traditional rehabilitation, maintaining a 1:1 patient-to-therapist ratio, targeting the same domains. Patients will undergo a total of 25 sessions, each lasting 45 minutes, with a frequency of 5 times a week, for inpatients; and 3 times a week, for outpatients. The primary endpoint is to demonstrate non-inferiority in the recovery of the activities of daily living as measured by the modified Barthel Index. If non-inferiority is established, the study will then evaluate the superiority of RADTs in the recovery of the activities of daily living. Secondary endpoints include improvements in upper and lower limb function, balance, cognitive function, and, according to the ICF, in the body functions, activities, and participation domains. Additional analyses will cover neurophysiological assessments of neural plasticity, as well as biochemical, and genetic evaluations. Upper limb dexterity and gait recovery rates during treatment will be monitored. The study will also evaluate daily activities and quality of life during a six-month follow-up period post-treatment. Acceptability and usability of integrated RADTs-based rehabilitation for patients, families, and healthcare providers, along with economic and organizational sustainability for patients, payers, and society, will also be assessed. Outcomes will be measured and analysed by blinded assessors.

**Ethics and dissemination:** This study was reviewed and approved by National Ethics Committee for clinical trials of Public Research Bodies (EPR) and other National Public Institutions (CEN). The results will be disseminated through scientific articles in peer-reviewed journals and presentations at both national and international conferences.

**Trial registration number:** NCT06547827

**Strengths and limitations of this study:**

• The effectiveness of robotics and allied technologies will be evaluated using a novel approach involving a multimodal intervention. This approach addresses all impaired functions to holistically enhance activities of daily living, which represent the patient’s most important needs.
• In addition to clinical scales, neurophysiological and biochemical data will be considered for a deeper understanding of the mechanisms underlying motor recovery.
• Detailed patient profiling will enable the creation of predictive models to identify subjects who may respond better to treatment integrated with robotics and allied technologies.
• The economic analysis will allow us to assess the sustainability of a new organizational model based on the use of robotics and allied technologies, where two physiotherapists supervise a group of patients simultaneously.
• A study limitation is the use of different technologies across research centres. However, this reflects real-world conditions. To address the inevitable variability, we clearly defined the functional domains for treatment with robotic or technological systems, focusing on the lower limbs, balance, upper limb segments, and cognitive functions. Moreover, to manage other possible sources of variability due to the number of centres involved, a consensus was reached about clear protocols for clinical evaluation and therapy administration to be shared in clinical practice by all the participating centres.

## Introduction

Within the rehabilitation field, Robots and allied digital technologies (RADTs) have been proposed as resources capable of revolutionising and enhancing treatment efficacy. Rehabilitation robots were primarily designed to amplify the dose of treatment (1), particularly in patients with severe motor deficits, and to alleviate the burden on physiotherapists (2). In traditional rehabilitation treatments, in fact, motor exercises are generally performed or assisted by physical therapists, mostly on one-on-one sessions, which have clear limitations related to the availability of time and human resources. These limitations may hamper access to care with a significant repercussions on treatment outcomes (3).

Rapid technological progress has led to the development of new devices that not only increase the quantity and intensity of treatment but also significantly enrich it. One crucial aspect is the use of sensory feedback, including visual, auditory, and tactile feedback, as well as virtual reality (4). These tools create a highly engaging "working" environment aimed at stimulating active and attentive participation from the patient, contributing also to maintaining the patient’s motivation.

Another important aspect is that robotic devices objectively measure the sensorimotor performance of the patients. In fact, robots are equipped with sensors that allow the collection of a vast amount of kinematic and kinetic data, providing concise indicators related to the biomechanics of patient’s movements (5,6). These data, if appropriately collected, provide an objective measurement of the patient’s status, enable tracking of their evolution over time, and allow for feedback regarding progress. One of the most promising applications of this data is the use of artificial intelligence algorithms, particularly machine learning, to assist rehabilitation professionals in data analysis and interpretation. This provides valuable information on patients’ recovery processes and supports the development of decision support systems that help clinicians select the most appropriate rehabilitative treatment for the patient, aligning with the perspective of precision medicine (7).

So far, the use of these algorithms has been limited to pilot projects, and the robotic devices currently available are not yet "intelligent machines", capable of adapting treatment to patient characteristics, unless they are used in adherence to protocols validated by expert personnel (8). This could be one reason for the non-superiority of robotic treatments over traditional ones (9,10) in some studies reported in the literature.

Indeed, numerous studies and meta-analyses (11–13) lead to inconclusive results regarding the effectiveness of robotics and technology in treating patients after stroke. This is due to multiple factors, particularly the considerable heterogeneity of available studies in terms of treatment duration, session frequency, and specific treatment modalities. Additionally, the limited number of treated patients represents a significant obstacle to the overall evaluation of effectiveness.

A major limitation of current scientific studies on post-stroke patients is that nearly all trials have utilized a single device, focusing solely on the robotic rehabilitation of an isolated body segment or function. This narrow approach overlooks the critical need for a holistic rehabilitation strategy that addresses the full spectrum of impaired functions in stroke patients. Comprehensive rehabilitation is essential to optimize recovery and improve the overall quality of life for these individuals, as it ensures that all affected areas and functionalities are treated in an integrated manner. Consequently, there is a need to conduct pragmatic studies on large case series to comprehensively and accurately assess the effectiveness of rehabilitation assisted by RADTs compared to traditional rehabilitation, adopting a global approach oriented towards the recovery of all compromised sensorimotor and cognitive functions (14). Indeed, these functions are closely interconnected: the recovery of one component can significantly influence the recovery of others. For instance, the recovery of cognitive functions can improve the ability to learn strategies and new motor skills, while the recovery of sensory functions can favour the planning and execution of the motor program. Conversely, the recovery of sensorimotor functions can improve participation and autonomy, which in turn can positively influence cognitive functions. Post-stroke rehabilitation is indeed a complex process, involving the sensorimotor functions of the upper and lower limbs, cognitive functions, and language, impacting activities such as walking, balance, and autonomy. For this reason, it is essential to consider function recovery and, more generally, rehabilitation as a "*multimodal, person-centred, collaborative process, including interventions targeting a person’s capacity and/or contextual factors related to performance, with the goal of optimizing the functioning of persons with disability*" (15). As the final goal of rehabilitation is to improve daily living activities, the recovery of a single movement (e.g., reaching for an object) is relatively unimportant if it does not translate into skills and autonomy (grasping, manipulating, and using the object itself).

Another crucial aspect is that the effectiveness of integrated multimodal rehabilitation with robots and other technologies in stroke patients may depend not only on treatment (in terms of intensity, frequency, and duration of sessions) but also on the physiological characteristics of the patient undergoing the intervention. In this sense, it is crucial to investigate biological processes and factors influencing the sensorimotor and cognitive recovery of the patient, such as those related to neurophysiological processes of brain plasticity, genetic expression of neuromediators (16), biomarkers production of neuronal damage (17–19), and nutritional alterations experienced by stroke patients (20). Detailed patient profiling can allow the creation of predictive models able to identify subjects who may respond better to RADTs-assisted treatment, making possible the development of targeted and personalized rehabilitation approaches.

Although not a primary target of the study, the collected data will enable further legal-ethical analysis to better understand the agency issues related to “delegating” professional treatment of patients. We hope that data can offer insights into device policy considerations able to identify criteria to authorize delegations of professional duties in line with existing legislations and ethical values.

In a real case scenario, along with proof of effectiveness, the economic evaluation and sustainability of robotic rehabilitation play a crucial role. To this aim, the study protocol will leverage Health Technology Assessment (HTA) (21). In a HTA framework, it is essential to realise the costs associated with robotic rehabilitation, including acquisition, maintenance, and staff training costs, and to compare them with the clinical benefits achieved, such as improvements in motor skills and patient quality of life. An accurate economic evaluation allows us to determine whether robotic rehabilitation represents an efficient use of healthcare resources while ensuring the long-term sustainability of RADTs-assisted treatment within the national healthcare system. Such considerations will be integrated with the legal and ethical ones strengthening eventual policy indications. Furthermore, a thorough sustainability analysis can identify potential areas for improvement and optimization, increasing the chance that a larger number of patients benefit from technology, and enhancing health administrators’ decision-making around the acquisition and implementation of robotics in their specific clinical setting (22–24).

Based on the above considerations, this paper describes the design of a pragmatic trial that will be carried out on a large sample of patients who have experienced a stroke within the six months prior to enrolment. Pragmatic trials aim at evaluating the effectiveness of an intervention in scenarios that mainly reflect clinical practices, in contrast with explanatory trials, whose aim is to evaluate the efficacy, i.e. the capacity to produce a desired effect in expert hands under ideal circumstances (25). As a matter of fact, while explanatory trials answer the question “Can an intervention work?”, a pragmatic trial focuses on the question “Does the intervention work?” in real-world clinical practice. The goal of the trial is to evaluate the effectiveness and sustainability of a multimodal treatment using robotics and advanced technologies compared to traditional multimodal treatment, in the recovery of daily living activities. Specifically, we aim to verify the non-inferiority of the robotic treatment, when compared to traditional interventions. We decided that a non-inferiority study was appropriate for the primary outcome, since there are no evident reasons so far for a robotic treatment to be superior to one-to-one traditional treatment, when the two treatments are administered with the same intensity. However, we are confident that this superiority may emerge, for example, for reasons related to patient engagement and compliance, and in any case, the technology might reduce healthcare costs, by allowing a lower therapist-to-patients ratio.

This pragmatic trial is part of Mission 1 on the clinical translation of robotics into rehabilitation, under the Fit for Medical Robotics (Fit4MedRob) Initiative, funded by the Ministry of University and Research of Italy.

## Methods and analysis

### Study objectives

#### Primary Objective

To demonstrate, in a population of subacute stroke patients, the non-inferiority of a rehabilitation treatment integrated with RADTs, compared to traditional rehabilitative treatment, in the recovery of activities of daily living.

#### Secondary Objectives

- To demonstrate the superiority of rehabilitative treatment integrated with RADTs compared to traditional rehabilitation treatment in the recovery of activities of daily living, should non-inferiority be demonstrated;
- To compare the improvements between the two groups in all targeted domains (upper limb, lower limb, balance, cognitive functions), and in accordance with the International Classification of Functioning, Disability, and Health (ICF) (26);
- To analyse the neurophysiological parameters and factors involved in neuroplasticity processes;
- To compare the time pattern of manual dexterity and walking performance recovery in the two groups;
- To assess the effects of the rehabilitation treatment in terms of daily life activities and quality of life through medium-term follow-up;
- To evaluate the acceptability and usability of the rehabilitative treatment integrated with RADTs for patients, their families, and healthcare practitioners;
- To create a model capable of predicting the effectiveness of robotic and technological treatment in post-stroke patients;
- To assess the economic sustainability of the rehabilitative treatment integrated with RADTs for the patient, payer, and society, through the creation of a model for the assessment and prediction of cost-effectiveness and cost-utility. Additionally, a Budget Impact Analysis will be performed from the perspective of the national healthcare system. To deal with uncertainty related to the values of the model parameters, each analysis will be accompanied by multiparametric sensitivity analyses.

### Study design

This study is a multicentre, multimodal, randomized, controlled, parallel (1:1), evaluator-blinded, pragmatic interventional study. Patients will be randomised to receive either a rehabilitation assisted by RADTs or a traditional rehabilitation treatment. The study is presented following to the Standard Protocol Items: Recommendations for Interventional Trials (SPIRIT) (27). Figure 1 summarises the study protocol, while Table 1 shows the SPIRIT schedule of enrolment, interventions and assessments. The study is registered on ClinicalTrials.gov (NCT06547827).

**Figure 1.**
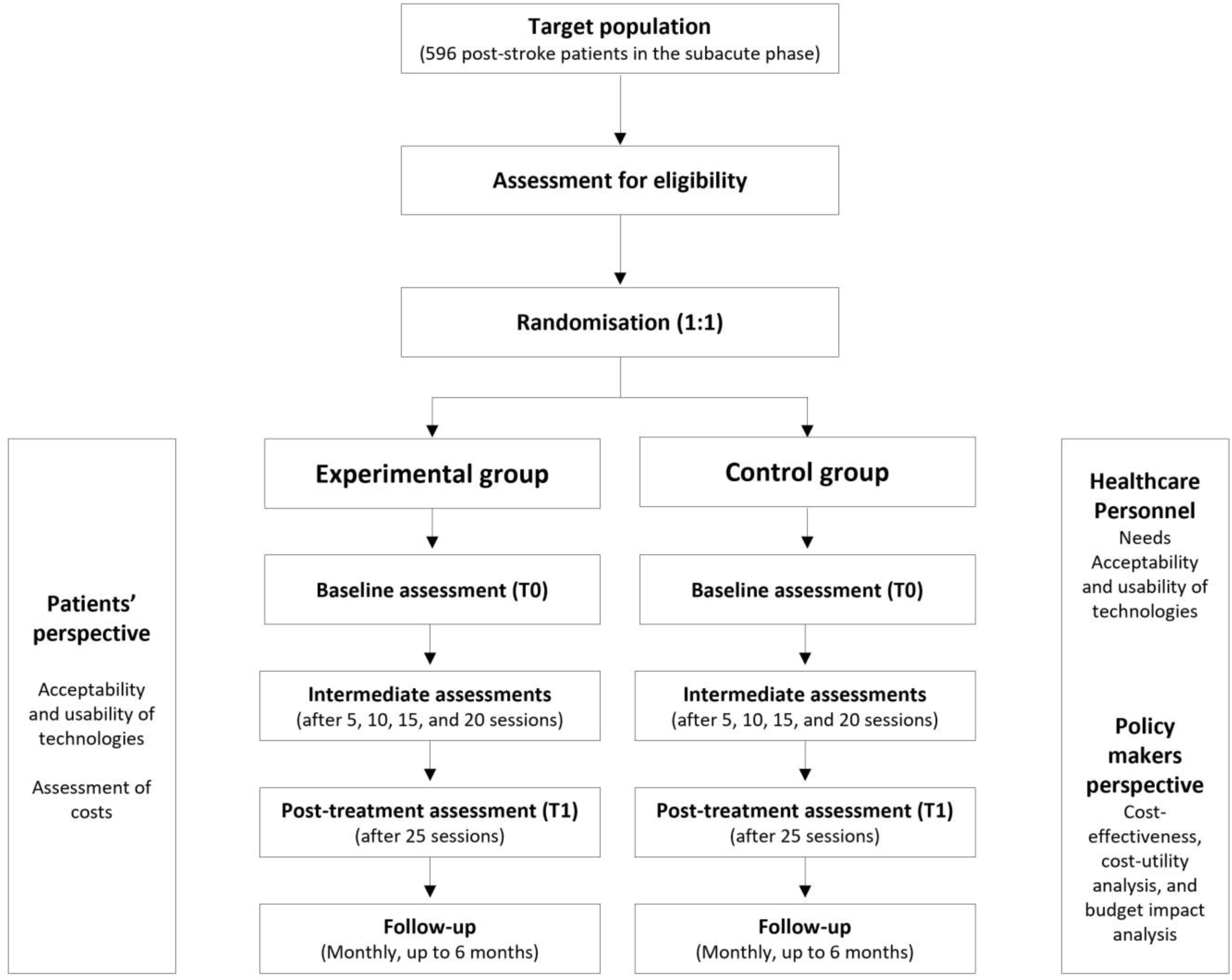
The STROKEFIT4 trial flow-chart.

**Table 1.**
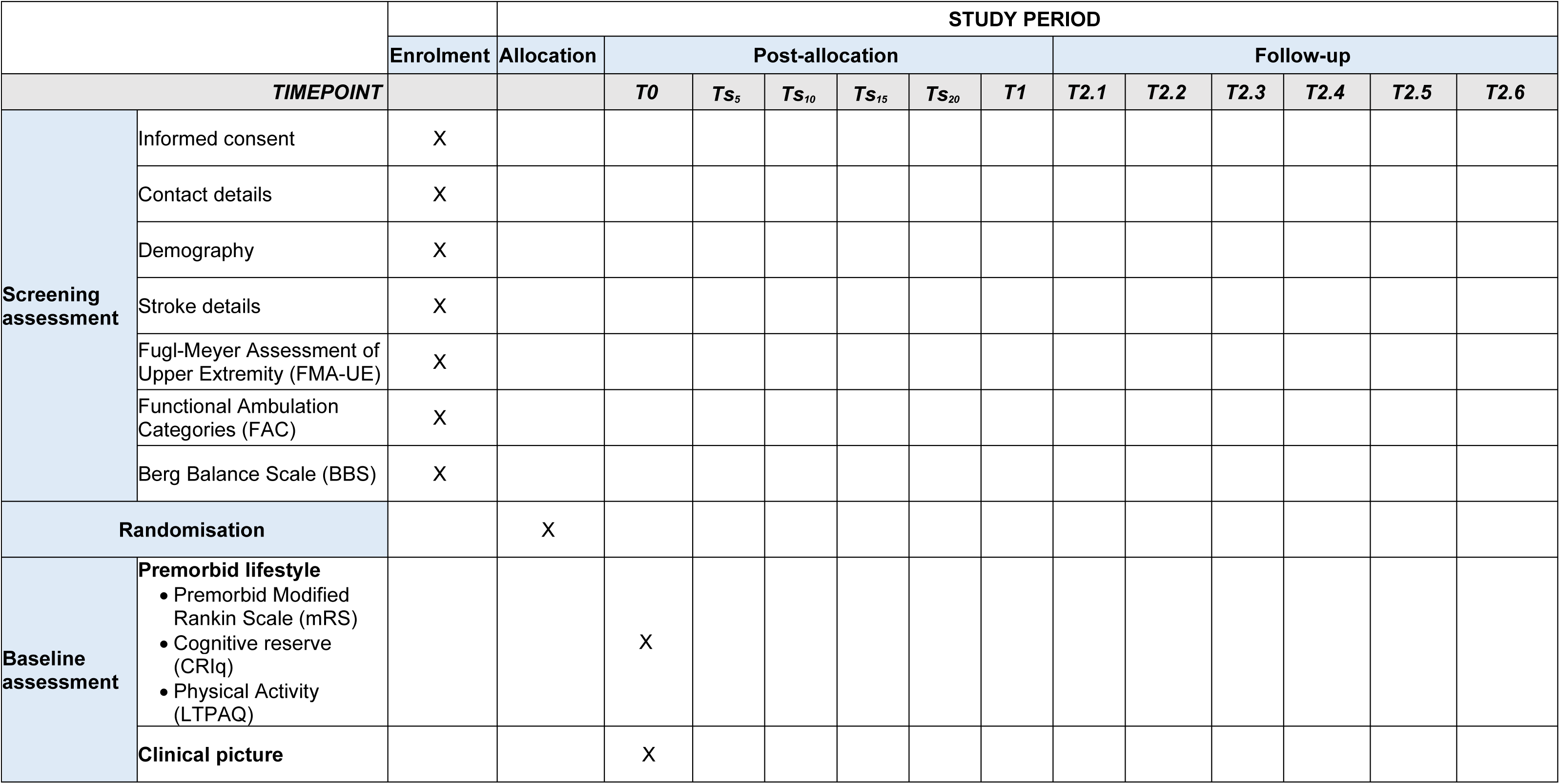
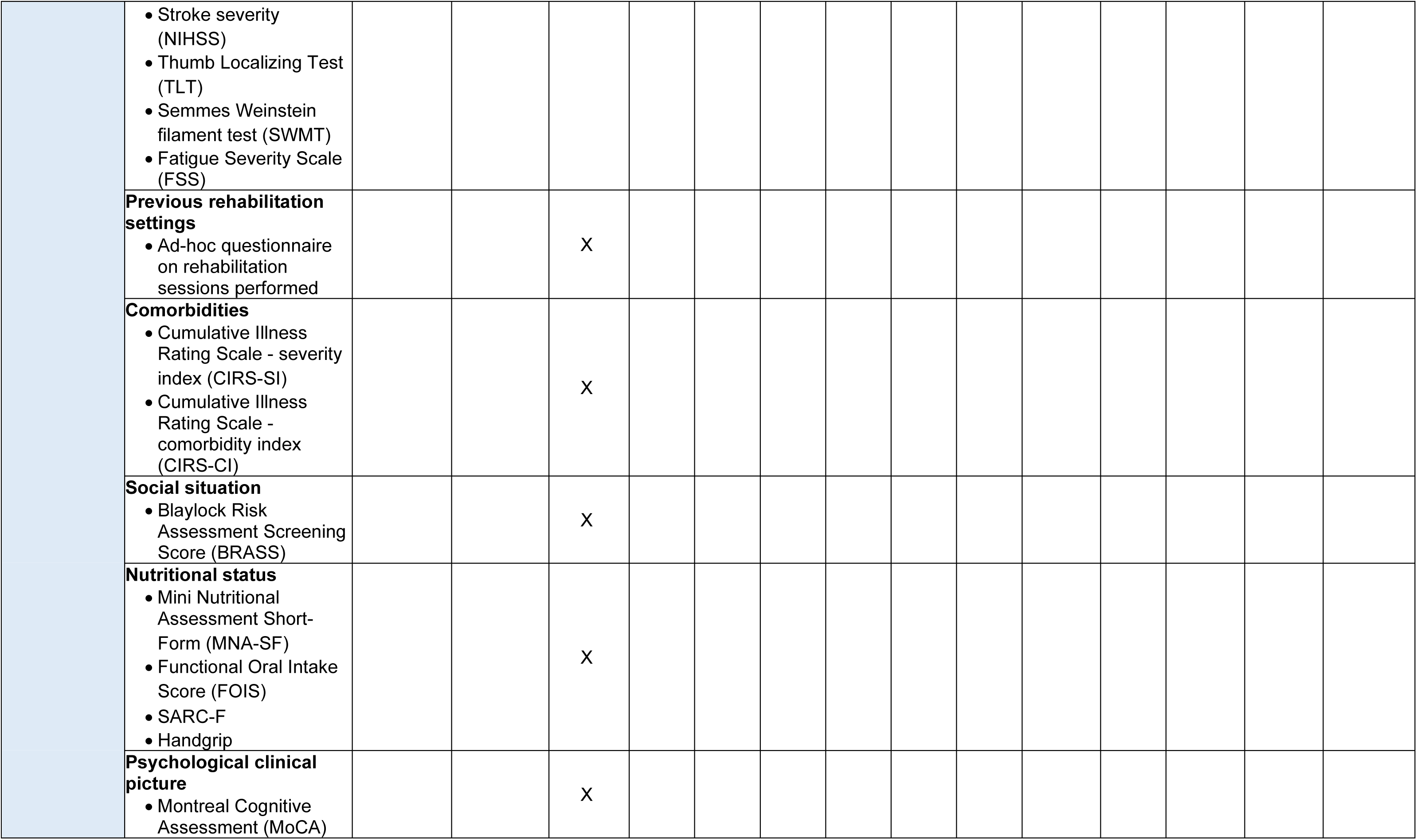
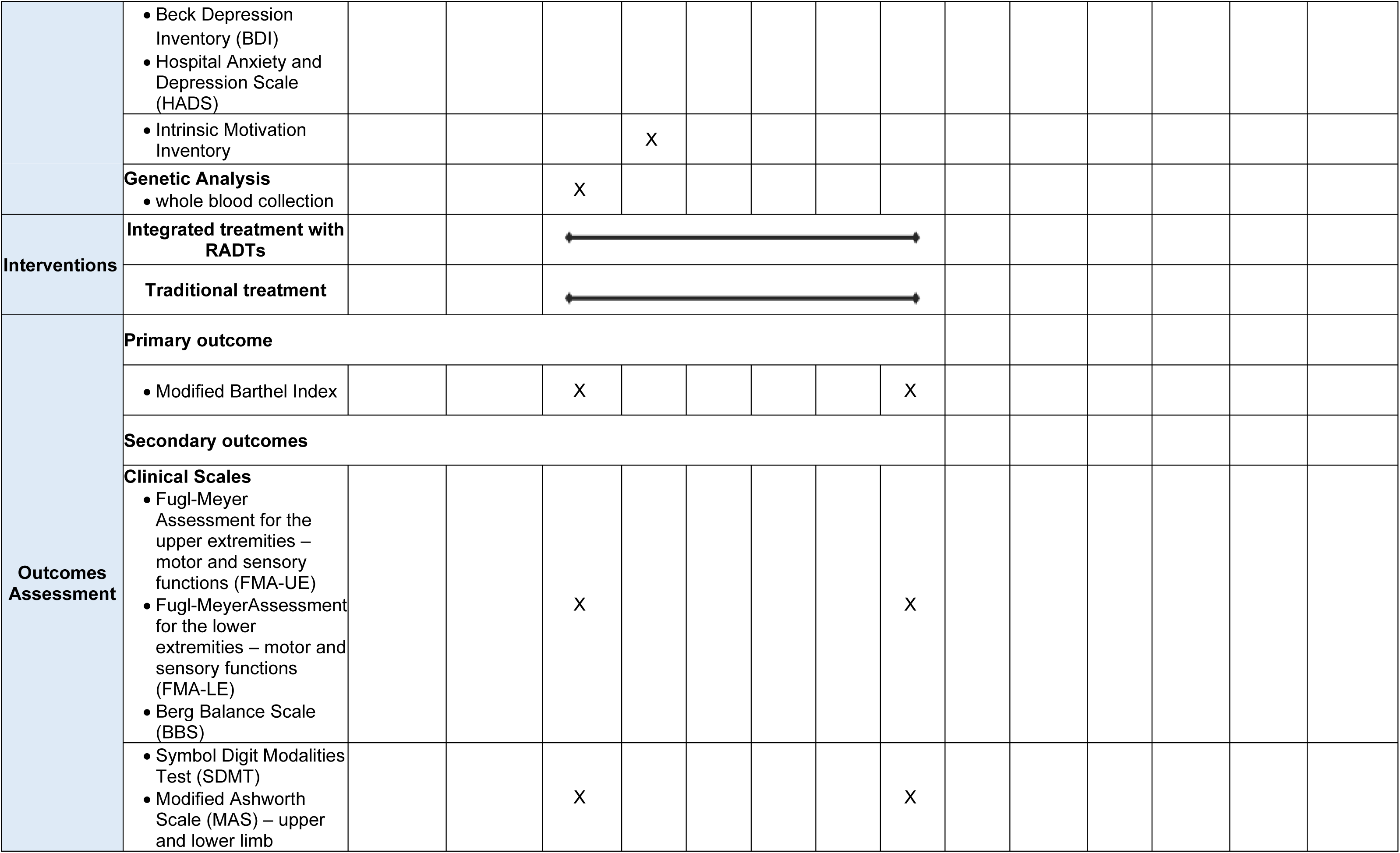
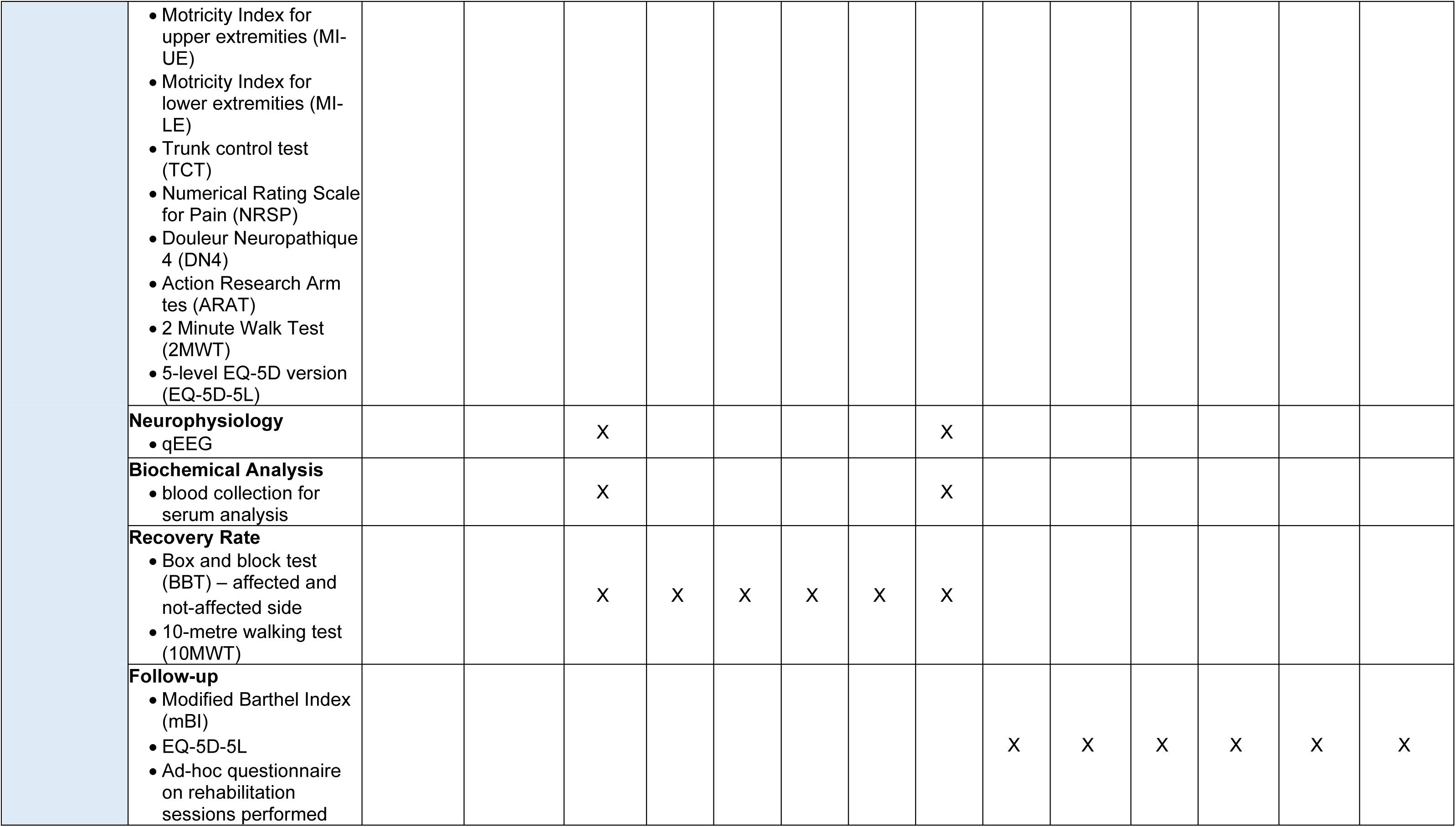
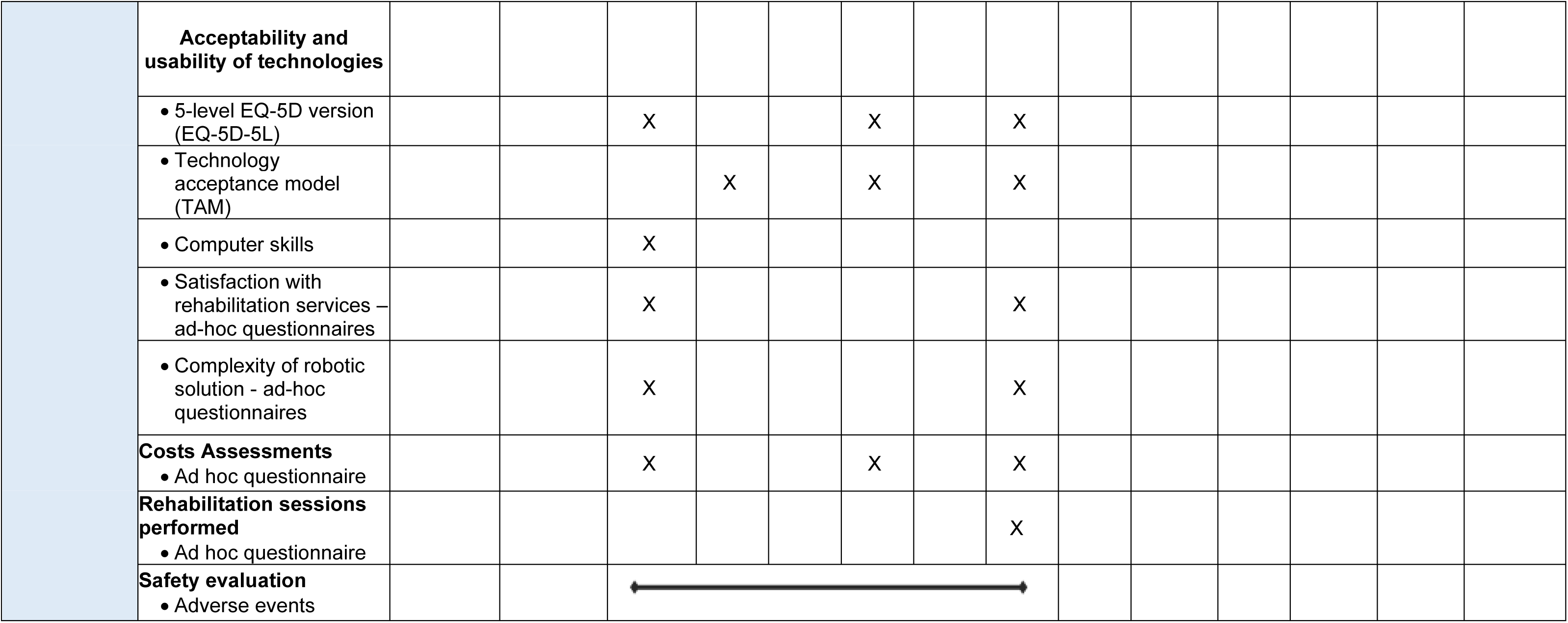
Standard Protocol Items: Recommendations for Interventional Trials (SPIRIT) schedule of enrolment, interventions and assessments. T0: baseline assessment (pre-intervention phase); T_is_: intermediate assessment (after the i-th rehabilitation session); T1: post-treatment assessment; T2.x: follow-up assessment (x months after T1)

### Study setting

The study will be conducted by four main clinical partners in several study centres providing inpatient and/or outpatient services, for a total of 13 recruitment sites. Participating centres are:

1. Don Carlo Gnocchi Foundation ONLUS, with the following six centres:

• Roma (RM), Centro Santa Maria della Provvidenza,
• Milano (MI), IRCCS Santa Maria Nascente,
• Sant’Angelo dei Lombardi (AV), Polo Specialistico Riabilitativo,
• Salerno (SA), Centro Santa Maria al Mare,
• Acerenza (PZ), Centro "Gala",
• Tricarico (MT), Polo specialistico riabilitativo;
2. IRCCS Mondino Foundation, one centre: Pavia (PV);
3. IRCCS Scientific Clinical Institutes Maugeri, with the following five centres:

• IRCCS Bari (BA),
• IRCCS Telese (BN),
• IRCCS Milano (MI),
• IRCCS Pavia (PV),
• IRCCS Montescano (PV);
4. IRCCS Ospedale Policlinico San Martino, one centre: Genova (GE).

### Study population

In this study, 596 stroke patients (ischemic or haemorrhagic) in the subacute phase (28), fulfilling the eligibility criteria that follow, will be enrolled.

#### Inclusion criteria

• First-ever diagnosis of ischemic or haemorrhagic stroke confirmed by Computed Tomography or Magnetic Resonance Imaging;
• · Age 18 years and over;
• Time since stroke equal to or less than 6 months;
• Mild to severe impairment of the upper limb (motor section of the *Fugl-Meyer Assessment of Upper Extremity* (29) ≤ 58) and/or mild to severe impairment of the lower limb (score on the *Functional Ambulation Categories* scale (30) ≤ 4) and/or mild to severe impairment of balance (*Berg Balance Scale* (31) ≤ 45);
• Clinical stability allowing transfer to the gym and execution of the planned treatments.

#### Exclusion criteria

- Clinical instability;
- Behavioural/cognitive disorders preventing adequate patient compliance with both traditional and robotic rehabilitation treatment (severe cognitive deficit, Montreal Cognitive Assessment (32) <10);
- Rigidity or hypertonia (*Modified Ashworth Scale* (33) = 4) in the plegic/paretic limb;
- Serious uncorrectable visual impairments preventing the patient from performing treatment with technological and/or robotic devices;
- Pregnant women;
- Refusal to sign the informed consent.

### Recruitment

Patients will be recruited from those attending rehabilitation at the aforementioned clinical centres, either as inpatients or outpatients. Enrolment will be competitive, with an expected number of at least 30 patients enrolled in each site. Specifically, individuals who had a verified diagnosis of stroke by magnetic resonance imaging (MRI) or computed tomography (CT) neuroimaging, will be identified by healthcare professionals and screened for eligibility. Once the compliance with the inclusion and exclusion criteria of the study is verified, the principal investigator at each recruitment centre, or individuals appointed by them, will be responsible for obtaining informed consent from patients after adequately informing them about the purposes, methods, expected benefits, and foreseeable risks of the study. During this time, the investigator will be available to answer questions and provide clarifications to ensure a proper understanding of the study.

The information sheet contains details regarding the purpose, methods, examinations, and assessments that the participant will undergo, any potential risks associated with them, and the procedures for pseudonymization that will be applied to the collected data.

Each study centre will maintain a screening log, which reports information regarding all inpatients and outpatients evaluated for potential inclusion in the study, as well as their subsequent inclusion or exclusion.

### Baseline assessment

Once written informed consent is obtained, a baseline assessment is performed by each study principal investigator, or delegated personnel.

#### Demographic and clinical characteristics

Concerning patient’s demographic and clinical characteristics, the following data will be collected:

• Demographic:

◦ Age
◦ Gender
◦ Handedness
◦ Height
◦ Weight
◦ BMI
◦ Smoker
◦ Years of education
◦ Current rehabilitation setting
• Past medical history;
• Acute event data;
• Medications taken;
• Premorbid lifestyle:

◦ Premorbid Modified Rankin Scale (34)
◦ Cognitive reserve (Cognitive Reserve Index questionnaire (35))
◦ Physical activity (Lifetime Physical Activity Questionnaire (36)
- Clinical picture:

◦ National Institutes of Health Stroke Scale (37)
◦ Thumb Localizing Test (38)
◦ Semmes Weinstein monofilament test (38)
◦ Fatigue Severity Scale (39)
• Previous rehabilitation settings:

◦ Ad-hoc questionnaire
• Comorbidities:

◦ Cumulative Illness Rating Scale (40)
• Social situation:

◦ Blaylock Risk Assessment Screening Score (41)
• Nutritional clinical picture:

◦ Handgrip test (affected and not affected side) (42)
◦ Mini Nutritional Assessment – Short Form (43)
◦ Functional Oral Intake Score (44)
◦ SARC-F (45)
• Psychological clinical picture:

◦ Montreal Cognitive Assessment (32)
◦ Beck Depression Inventory (46)
◦ Hospital Anxiety and Depression Scale (47)
◦ Intrinsic Motivation Inventory (48)

#### Genetic analyses

• The Brain-derived neurotrophic factor (BDNF) rs6265 genotyping of patients will be evaluated according to the following procedure (16):

◦ Whole blood samples will be collected at T0 (2 X 4 mL purple cap tubes, EDTA) and frozen (-20°C or -80°C) until shipment/analysis;
◦ DNA extraction of whole blood samples will be done with a research extraction kit (Zymo Research, Irvine, CA, USA);
◦ The BDNF rs6265 genotyping will be performed using a polymerase chain reaction (PCR) combined with restriction enzyme digestion (HpyCH4IV enzyme). The electrophoresis resolution of fragments will detect, for each patient, the presence of a valine (Val) to methionine (Met) substitution at codon 66 (Val66Met). Patients will be then identified as “non-carrier” of substitution (homozygous *GG*), and “carrier” of A substitution (Met protein replacement at codon 66, as heterozygous *AG* or homozygous *AA*).

### Randomisation

Patients will be randomly assigned either to a robotic-technological rehabilitation treatment or to a traditional rehabilitation treatment. The randomisation will be centralised, overseen by an experimenter not directly involved in patient recruitment, treatment, or evaluation, and will be stratified by clinical centre, time since stroke (inferior to 3 months; between 3 and 6 months), and clinical setting (inpatient or outpatient). Randomisation lists will be generated using the National Cancer Institute Clinical Trial Randomization Tool (https://ctrandomization.cancer.gov/).

### Interventions – common characteristics

All randomized subjects, in accordance with the ICF (26), will undergo rehabilitation for the following functions and activities, tailored to each patient according to the guidelines (49–54):

1. Sensorimotor Functions

a) Neuro-musculoskeletal and movement-related functions:

• · Mobility and stability of joint functions in one or more joints of the upper limb, lower limb, and spine;
• · Muscle tone and strength;
• · Movement functions.
b) Sensory and pain functions;
c) Exercise tolerance function (physical endurance, aerobic capacity, and fatigue resistance).
2. Specific Mental Functions (attention, memory, perceptual functions, higher-level cognitive functions, language and calculation mental functions, self- and time-experience).
3. Activities and Participation: self-care and daily life activities (washing, grooming, dressing, eating, etc.).

#### Rehabilitation of Sensorimotor Functions

A) Neuro-musculoskeletal and Movement-related Functions The neuro-musculoskeletal and movement-related functions of the plegic/paretic upper limb in the proximal (shoulder), intermediate (elbow), and distal (hand) districts will be rehabilitated through:

• Passive, active, and active-assisted exercises involving the three major joints of the limb in their degrees of freedom, exercises for maintaining reciprocal joint relationships, exercises facilitating scapula and carpal bone movement;
• Muscle tone control exercises, incremental muscle recruitment exercises, and muscle endurance exercises;
• Coordination exercises of voluntary movements, spasticity inhibition exercises, management exercises of muscle stiffness. by muscle and joint stretching

Objectives of these exercises will be (i) facilitation of sensory-motor reorganization of the upper limb to improve strength, motor function, and performance; (ii) improvement of range of motion of the upper limb; (iii) prevention of complications from disuse (musculo-tendinous, capsulo-ligamentous and joint retractions). The neuro-musculoskeletal and movement-related functions of the plegic/paretic lower limb in the proximal (hip), intermediate (knee), and distal (ankle) districts will be rehabilitated through:

• Passive, active, and active-assisted exercises involving the three major joints of the limb in their degrees of freedom, exercises for maintaining reciprocal joint relationships, exercises facilitating pelvic and tarsal bone movement;
• Muscle tone control exercises, incremental muscle recruitment exercises, and muscle endurance exercises;
• Control of reactions (postural, body straightening, body adjustment, balance, support, and fall defence), coordination exercises of voluntary movements, spasticity inhibition exercises, management exercises of muscle stiffness by muscle and joint stretching, and gait training.

Objectives of these exercises will be: (i) facilitation of sensory-motor reorganization of the lower limb to improve strength, motor function, and performance; (ii) improvement of joint mobility of the lower limb; (iii) improvement of static and dynamic balance; (iv) relearning of movement patterns associated with fractional step and full-cycle walking; (v) prevention of musculotendinous and capsulo-ligamentous retractions, and joint rigidity by disuse.

B) Sensory function Sensory function will be rehabilitated through:

• Targeted proprioceptive exercises for the proprioceptive and kinaesthetic functions of the plegic/paretic upper and lower limbs;
• Proprioception exercises related to sitting and standing positions, static and dynamic balance exercises (in terms of displacements, directional changes, and speed in monopodalic and bipodalic conditions);
• Reduction and control of pain sensation.

Objectives of these exercises will be: (i) reorganization of body position perception and spatial sense; (ii) stimulation of peripheral receptors to correct and improve physiological movement performance; (iii) promotion of facilitatory movement management in terms of performance and ergonomics.

C) Exercise Tolerance Functions Exercise tolerance functions, including physical endurance, aerobic capacity, and reliability, will be treated with:

• General physical endurance exercises;
• Gradual, progressive, and prolonged aerobic exercises;
• Fatigue control and management at varying effort levels.

Objectives of these exercises will be (i) improvement of endurance; and (ii) reconditioning of cardiovascular and respiratory systems.

#### Rehabilitation of Specific Mental Functions

Specific mental functions (attention, memory, perceptual functions, higher-level cognitive functions, language and calculation mental functions, and self- and time-experience functions) will be treated with:

• Attention training exercises (e.g., visual search exercises, barrage);
• Memory training exercises (e.g., repetition and delayed recall of word lists);
• Recognition and interpretation exercises of sensory stimuli;
• Exercises for higher-level cognitive functions (e.g., decision-making processes, planning, problem-solving, etc.);
• Exercises for language-specific mental functions (recognition and use of signs, symbols, and other language components);
• Exercises for specific mental functions of simple mathematical calculations and complex mathematical operations;
• Exercises for awareness of one’s identity in the reality of one’s environment and time.

Objectives of these exercises will be (i) improvement in patient orientation in spatial-temporal dimensions and patient’s reality orientation; (ii) increase in short-term memory spans; (iii) improvement in attention times and concentration capacity on a task; (iv) improvement in selective and sustained attention; (v) improvement in language comprehension and production skills; (vi) maintenance of acquired and residual skills in daily life.

#### Rehabilitation of Activities and Participation, Self-care, and Daily Life Activities

Daily life activities (washing, grooming, dressing, eating, etc.) and participation will be treated with:

• Exercises of learning and application of learned knowledge and problem-solving;
• Execution of single or multiple tasks and routine organization exercises;
• Mobility (postural passages; transfers; object transportation and movement; task-oriented and manipulation exercises; walking and moving in different places and using devices);
• Activities aimed at self-care, washing and drying, dressing, and eating;
• Performance of domestic and daily activities;
• Reintegration into social life.

Objectives of these exercises will be (i) improvement of executive functions (problem-solving) with compensation strategies; (ii) safety protection (risk of falling); (iii) improvement of movement control and precision; (iv) identification and elimination of environmental barriers; (v) improvement of physical, psychological, and social adaptation capabilities; (vi) identification of strategies aimed at carrying out daily life activities (personal care, work, school, and leisure time).

### Specific interventions

#### Experimental Group: Integrated Treatment with Robotic & Technological Devices

The experimental intervention involves administering a rehabilitative treatment, as described above, using (also) RADTs. Based on the equipment available at each centre, different devices may be used; however, harmonization of the treatments provided across all centres will be ensured by using, in each centre, devices that enable rehabilitation in the following domains:

A) Upper limb sensorimotor abilities;
B) Lower limb sensorimotor abilities and Gait;
C) Balance;
D) Cognitive abilities.

Specifically, for rehabilitating sensorimotor function of the upper limb in the proximal (shoulder), semi-distal (elbow), and distal (hand) regions, the following devices may be used:

• Planar end-effector robots for shoulder-elbow rehabilitation, or exoskeletons or electromechanical systems for shoulder, elbow, and wrist rehabilitation;
• End-effector robots or exoskeletons for the hand;
• Sensor-based devices for comprehensive upper limb treatment.

For rehabilitating sensorimotor function of the lower limb, the following ones will be considered:

• End-effector robots or exoskeletons (overground and non-overground) for the lower limb;
• Treadmills with body-weight support systems.

Balance training under static and dynamic conditions will be ensured by:

• Robotic and digital stabilometric platforms;
• Sensor-based systems.

For cognitive functions, digital systems, sensor-based devices, and virtual reality programs will be used to train memory, attention, visuospatial skills, and executive functions.

The treatment will be conducted within a gym equipped with devices enabling treatment of all identified domains, where two physiotherapists will supervise a group of patients ranging from 4 to 6, depending on the severity of their clinical condition.

All devices used in the study must already have a CE mark for medical devices and be used according to the manufacturer’s intended use.

Information on the experimental intervention with RADTs were reported following the *Template for Intervention Description and Replication (TIDieR) Checklist* (55).

#### Control Group: Traditional Treatment

In the control group, subjects will undergo a standard traditional rehabilitation program without the use of RADTs, focusing on activities and objectives mentioned in the section “Interventions”, using traditional methods of physiotherapy and cognitive rehabilitation. As in usual clinical practice, a physiotherapist-to-patient ratio of 1:1 is provided for the treatment in all participating centres.

#### Intervention Schedule

Patients in the experimental group will undergo a total of 25 robot-assisted sessions, each lasting 45 minutes, with the following frequency:

• 5 times a week for 5 weeks, for inpatients;
• 3 times a week for 8.3 weeks, for outpatients.

The difference in treatment frequency between the two settings is related to current regulations and the difficulty as an outpatient to travel to urban rehabilitation facilities on a daily basis. This approach ensures that patients receive the same number of rehabilitation sessions and are able to attend all scheduled sessions, thus reducing the risk of potential dropouts.

In addition, according to current Italian regulations, which require providing inpatients with 3 sessions/day of treatment for 6 days/week, all inpatients (also those randomised in the intervention arm) will undergo, in addition to the 25 sessions, traditional rehabilitation sessions (6 times/week), lasting 45 minutes. To avoid the possibility of performance bias, the therapists who will treat subjects in RADT-assisted treatment will be different from therapists who will treat the same subjects traditionally.

Moreover, according to the medical prescription, all subjects (inpatients and outpatients) will undergo one session/day of occupational and/or speech and swallowing therapy, based on their needs.

All rehabilitation sessions including type (neuromotor, cognitive, occupational therapy, speech therapy), method (robotic or conventional), duration in minutes, and possible issues arising, will be recorded daily by the therapists (session diary).

### Intervention Adherence

High adherence to the study is expected, as patients will undergo treatment during the period of admission to the rehabilitation facilities (albeit in different settings, inpatient or outpatient, depending on the severity of the clinical condition) in accordance with current regulations. To ensure the quality and consistency of the intervention, adherence will be carefully assessed by an independent team of clinical operators, who will regularly monitor the session diary. If needed, targeted strategies will be implemented to maintain and improve adherence, including periodic training sessions for the involved staff and providing timely feedback to the therapists involved in treatment in both study groups.

### Outcome assessments

#### Primary Endpoint

The primary endpoint of the study will be the change in the modified Barthel Index (35) between the beginning and the end of the treatment (T0-T1).

#### Secondary Endpoint

The study includes numerous secondary outcomes, aimed at capturing changes in patients who may have very different sensorimotor functions and performances, considering that we are examining patients within the first 6 months after the acute event and in different settings.

1 Treatment Effects, measured by clinical scales at the beginning (T0) and end of the treatment (T1)

• Fugl-Meyer Assessment for the upper extremities (upper limb sensorimotor domain) (29);
• Fugl-Meyer Assessment for the lower extremities (lower limb sensorimotor domain) (29);
• Berg Balance Scale (balance domain) (31);
• Symbol Digit Modalities Test (cognitive domain) (56);
• scales in accordance with the domains of the ICF:

• Body function

◦ Modified Ashworth Scale (33);
◦ Motricity Index (upper and lower extremities and trunk) (57) ;
◦ Numerical Rating Scale for Pain (58) ;
◦ Douleur Neuropathique 4 (59);
• Activities

◦ Action Research Arm test (60);
◦ 2 Minute Walk Test (61);
• Participation

◦ EQ-5D-5L (62).
II. Treatment Effects – Neurophysiology At the beginning (T0) and end of the treatment (T1), all patients will undergo a neurophysiological evaluation through high-density resting electroencephalography (EEG) to study the neurophysiological recovery processes. Both groups of patients will undergo high-density EEG recording, using a 64-channel HD-EEG system. The signals will be acquired using a cap with 64 electrodes positioned according to the international 10/20 montage. Contact impedance will be maintained below 5KΩ. Recordings will be made for 10 minutes with eyes open and 10 minutes with eyes closed, with the subject relaxed and in a comfortable supine position. Specifically, from the recorded signals, the Brain Symmetry Index (BSI) (63,64), a quantitative electroencephalographic (qEEG) measure used to assess the degree of symmetry in brain activity between the two hemispheres, and the Delta/Alpha Ratio (DAR) (65), a qEEG measure used to assess the balance between slow-wave (delta) and fast-wave (alpha) brain activity, will be obtained.
III. Treatment Effects – Biochemical Analysis In all patients, changes in the levels of BDNF, neurofilament light (NfL), and glial fibrillary acidic protein (GFAP), which are factors involved in neuroplasticity and tissue damage, will be evaluated. Specifically, at T0 and T1, blood samples will be collected in the morning after overnight fasting (two 6mL yellow cap tubes, for biochemical analyses); tubes will be then centrifuged at 3000 rpm for 15 minutes, aliquoted (each aliquot will have a volume of 500 microliters), and frozen (-20°C or -80°C) until shipment/analysis. Sera samples will be thawed just before the assay and will be analysed on an automated immunoassay system (Ella Simple Plex, Biotechne) to measure serum levels of BDNF, NfL and GFAP.
IV. Time pattern of recovery during Rehabilitation The time pattern of recovery during treatment will be assessed by administering the following tests every 5 treatment sessions:

• Box and block test (66);
• 10-meter walk test (67).

During the execution of the motor tasks required by the two tests, where possible, patients will be instrumented with inertial sensors and surface electromyography to obtain quantitative information on their motor performance.

V. Follow-up

At the end of treatment, the patient will be contacted every month for the following 6 months, and the following questionnaires will be administered:

• Ad-hoc questionnaire on the rehabilitation activity performed (type of rehabilitation setting, number of sessions, frequency, duration, type of treatment, etc.);
• Modified Barthel Index;
• EQ-5D-5L.

In case the patient is unable to reach the clinical centre, the questionnaires will be administered by phone or other remote connections. As a matter of fact, those scales can be administered remotely and allow the evaluation of general aspects related to skills and participation.

### Costs, Sustainability and Acceptability Assessments

The sustainability of the proposed solutions will be analysed through the comparison of their estimated costs and effects. We will employ the following analyses: cost-effectiveness analysis, cost-utility analysis, and budget impact analysis.

Cost-effectiveness analysis (CEA) will compare the costs and clinical outcomes of the alternative rehabilitation models being studied. The cost-effectiveness ratio (ICER) will be calculated as the difference in costs between the alternative rehabilitation models tested in the clinical study and the difference in clinical outcomes (objective measures by physical units). The ICER represents the additional cost incurred to achieve an incremental unit of effectiveness, that will be assessed considering primary and secondary outcomes. Cost-utility analysis (CUA) will assess and compare the efficiency of alternative rehabilitation interventions by measuring health outcomes in terms of utility, a measure of general well-being or quality of life (subjective measures), relative to the costs incurred to achieve these outcomes. Utilities will be evaluated using standardized measures based on quality of life. Each health condition identified through generic health-related quality of life questionnaires (EQ-5D-DL) will be associated with available utility scores specific to Italy (68). The incremental cost-utility ratio (ICUR) will serve as the key metric, calculated as the ratio between differences in costs and differences in utility across the analysis groups.

From the perspective of the payer, the expected outcome of the budget impact analysis is the assessment of the financial impact (differences in direct health costs) over a 3–5-year period for the diffusion of a mix of robotic-based rehabilitations, compared to traditional rehabilitation. The financial impact will be assessed by considering the direct health costs associated with the different rehabilitation regimes, varying projections of robotic market uptake, and the degrees of substitutability between the compared rehabilitation regimes. Sensitivity analysis will be performed to assess the robustness and reliability of the results, evaluating how small variations in the variables considered affect the synthesis outcomes (ICER, ICUR, and budget impact).

#### Questionnaire for patients

Several questionnaires will be provided to patients, to analyse:

• Acceptability and usability

◦ Technology Acceptance Model (69)): after 5 sessions, after 15 sessions, at T1;
◦ Ad-hoc questionnaire for needs and available solutions and perception of their relative complexity: at T0 and T1;
• Satisfaction (ad-hoc questionnaire): at T0 and T1;
• Confidence and knowledge of ICT technologies (ad-hoc questionnaire): at T0;
• Costs (ad-hoc questionnaire): at T0, after 15 sessions, at T1;
• Quality of life (EQ-5D-5L (62)): at T0, after 15 sessions, at T1.

#### Questionnaires for operators

In addition to the study and control groups, the group of healthcare professionals involved in rehabilitation will also be included in the analysis, and they will be asked to respond to questionnaires regarding:

• Confidence and knowledge of ICT technologies: at the beginning of the study;
• Quality of work (Work-Related Quality Of Life scale, (70)): at the beginning of the study, 6 months after, and 12 months after;
• Technology Acceptance Model (69): at the beginning of the study, 6 months after, and 12 months after;
• Ad-hoc questionnaire for needs and available solutions and perception of their relative complexity: at the beginning of the study, 6 months after, and 12 months after;
• Satisfaction (ad-hoc questionnaire): at the beginning of the study, 6 months after, and 12 months after.

### Patient’s involvement in the study design

The study design was developed based on the needs identified through a survey conducted by the Fit4MedRob Consortium, targeting both patients and healthcare professionals. The survey revealed that many respondents experience simultaneous impairments across multiple functional domains, such as movement, posture and balance, upper limb function, cognitive abilities, self-care, and communication. Additionally, the survey highlighted economic concerns, with respondents indicating that treatments are only partially reimbursed by the National Health System, underscoring a significant financial burden on patients.

### Sample size

As previously reported (71,72), a change of 9.25 points in the 0-100 version of the mBI corresponds to a change of 1.85 points on the 0 to 20-point version of the BI, which has been shown to be clinically significantc (73). Thus, an MCID (minimal clinically important difference) of the mBI of 9.25 points can be considered. The sample size was thus determined considering the following, with respect to the primary outcome (the mBI):

• the non-inferiority of robotic treatment compared to traditional treatment;
• a non-inferiority margin of 5 points (about half of the MCID);
• a statistical power of 80%;
• a bilateral 95% confidence interval;
• a standard deviation of the primary outcome of 20 points.

Considering these parameters, a sample size of 506 patients (253 per group) is obtained. Additionally, considering a dropout rate of 15%, a total sample size of 596 patients is obtained. The same sample size is sufficient to also demonstrate the potential superiority of robotic treatment. Considering a significance level of 5%, a statistical power of 80%, an MCID of 9.25 points, a standard deviation of 20 points, and a dropout rate of 15%, a total of 178 patients are required to demonstrate superiority. All calculations were performed using the software G*Power v.3.1.9.7 (74).

### Blinding

Given the nature of the study, neither patients nor clinical investigators can be blinded to the treatment; however, evaluations will be conducted by evaluators unaware of the treatment group, and similarly, data analysis will be conducted blindly.

### Study withdrawal

Participants have the freedom to withdraw from the study at any point and for any reason, providing notification to the principal investigator of the study. In this case, no further data concerning the patient will be collected, and he/she may request the deletion of the data already collected. Current and future medical care at the corresponding clinical centre will not be affected by patients’ decisions, and the physicians will continue to treat him/her with due attention. Additionally, investigators retain the authority to remove participants from the study if they believe it is no longer beneficial for them to participate, such as due to unforeseen illness or adverse events.

### Statistical analysis

The collected data will be presented using descriptive statistics such as mean and standard deviation for numerical variables, and percentages for categorical variables.

An Intention-To-Treat (ITT) population will be defined to include all patients randomised to the experimental group or the control group. A Per-Protocol (PP) population will be defined to exclude randomised patients who were not eligible at baseline (i.e., patients inappropriately randomised) and those who did not receive the full treatment (due to treatment interruption for any reason).

#### Primary analysis

The primary endpoint of Barthel index variation at the end of treatment will be compared between treatment groups using a mixed-effects linear model with the clinical centre as a random effect to obtain a corrected estimate of variance and, therefore, a correct confidence interval for the difference between the changes in the two groups. Non-inferiority will be assessed at a significance level of 2.5% by estimating the effect with a bilateral 95% confidence interval (CI) and comparing the lower limit of the CI with the non-inferiority margin of -5 points. For the analysis of the primary outcome, the mixed-effects model will initially be applied to the PP population. The analyses will be completed according to the Intention-To-Treat strategy (75,76), including all randomized subjects, along with a sensitivity analysis.

The ITT population will be defined by assuming a follow-up, and therefore an evaluation of outcomes, on subjects who interrupt treatment. The analysis will be conducted assuming that missing data are “missing at random” (MAR): therefore, a mixed model for repeated measures will be applied, including all subjects in the analysis. With respect to the assumption of MAR, a sensitivity analysis will be performed considering scenarios deviating from it and involving one or both groups. Additionally, a sensitivity analysis will be used to assess additional adjustments for baseline Barthel index values.

If non-inferiority is concluded for the primary outcome, superiority can be evaluated without the need for correction for multiple comparisons.

Regarding the primary outcome, an analysis of planned subgroups will be conducted to assess the presence of treatment effect modifiers, extending the models to include interaction terms between these factors and treatment.

#### Secondary analyses

Continuous secondary outcomes will be analysed only in the ITT population with the same model specification as the primary outcome and reported as adjusted mean differences. All tests will be two-tailed with a significance level of 5% and effect estimates will have a 95% CI.

Categorical outcomes will be reported as proportions with 95% CI and compared using Pearson’s χ² test, or Fisher’s exact test and exact binomial CI in case of frequencies less than five in contingency tables.

For comparing the trend over time of assessments obtained through the Box and block test and the 10-meter walk test in the two treatment groups, mixed models for repeated measures will be applied. The temporal trend of assessments will be estimated and compared in the two groups by introducing interaction terms between time and treatment group into the models.

A similar analysis will be conducted to estimate the trend of parameters observed during follow-up. This analysis will include subjects who have completed the treatment, i.e., the PP population, which will be studied over the 10 months following the end of treatment.

Finally, potential prognostic factors will be explored by including them as covariates in mixed models for repeated measures. These models will have, as the response variable, the outcome of interest and, among the explanatory variables, the treatment group, the clinical centres, and the factors whose effect on the outcome is to be estimated, also analysing possible interaction effects with treatment.

### Safety evaluation

Given the nature and objectives of the study, there are no foreseeable adverse events related to the conduct of experimental activities. The only adverse effects to consider are related to the treatment that the involved patients will undergo. However, the likelihood of risks and side effects is low since no invasive treatments or therapies are planned. The devices are CE-marked and have been in use at some of the investigational centres since 2016, where they have been used to treat hundreds of patients and thousands of rehabilitation sessions. Furthermore, the treatment will always be supervised by experienced physiotherapists. In our experience and the literature, no adverse events have been reported. In any case, regarding any reportable events, the mode and timing of reporting will refer to the obligations imposed by EU Regulation 2017/745 (Article 80) and the MDCG Guideline 2020-10/1 ("Safety reporting in clinical investigations of medical devices under the Regulation (EU) 2017/745").

## Discussion

Robotics and technology in the field of rehabilitation have been proposed as tools that can enhance the effectiveness of treatments. Early rehabilitation robots were specifically designed to increase the quantity and intensity of therapy, particularly for patients with severe motor impairments, while also reducing the workload on physical therapists. Traditionally, rehabilitation treatments rely on clinical staff to either assist or directly facilitate patients’ movements, which presents limitations in terms of time and human resources, often impacting treatment outcomes. An equally important advantage of using robots is their ability to provide objective measurements of motor performance. Equipped with sensors, these devices can collect objective data, such as kinematic or dynamic metrics, and generate precise indicators related to the biomechanics of a patient’s movements. When properly collected, this data offers a reliable assessment of the patient’s condition and allows for tracking progress over time.

Despite the potential benefits of using robots and technology in rehabilitation, several barriers hinder their widespread adoption in clinical practice. Although these technologies show promise in enhancing the measurement, intensity, and personalization of rehabilitation treatments, their high costs—encompassing not only the purchase of the equipment but also the training required for practitioners—remain a significant challenge along with legal and ethical considerations. Regarding the effectiveness of robotics and technology for post-stroke patients, the scientific literature includes numerous meta-analyses. However, these studies often yield inconclusive results due to various factors, notably the substantial variability in treatment duration, session frequency, and specific therapeutic approaches across different studies. Additionally, the small sample sizes in many studies pose challenges for evaluating overall efficacy. This underscores the need for pragmatic studies on larger, real-world populations, using a consistent treatment and measurement model while maintaining clinical feasibility within current regulatory frameworks. It is therefore essential to assess both the effectiveness and sustainability of these technologies in clinical practice through large-scale pragmatic trials to generate robust findings. For this reason, we have designed a pragmatic trial involving a broad sample of post-stroke patients in the subacute phase (within six months of the acute event).

To our knowledge, this is the first trial in the literature that will study the effectiveness of a multimodal, multidomain rehabilitation based on the use of robotics and allied technologies on such a large sample of subacute phase stroke patients. As mentioned, rehabilitation after a stroke is a comprehensive process involving all impaired functional domains namely sensory-motor function of the upper limb (in the proximal and distal district), lower limb and gait, balance and cognitive functions. Today, robots and technologies that can rehabilitate all those four domains are available. Thus, unlike previous trials that evaluate the effect of a robot in one segment (upper limb or lower limb), this trial proposes a treatment model using a set of robots to treat up to four functions. In fact, our primary outcome is the patient’s ability in activities of daily living in which the use of the upper limb is fundamental both in its reaching component but also in the grasping of objects; similarly in the skills of daily living the function of gait and thus balance is fundamental; on the other hand, all these functions are strongly influenced by cognitive abilities (a patient with impaired attention and planning for example will have difficulty in performing complex movements).

Post-stroke rehabilitation is indeed a complex process, involving the sensorimotor functions of the upper and lower limbs, cognitive functions, and language, impacting activities such as walking, balance, and autonomy. We consider it crucial to adopt a global approach oriented towards the recovery of all compromised sensorimotor as well as cognitive functions (14) because all these functions are closely interconnected. We decided to use daily living abilities as the primary outcome because we believe it is the most important goal of recovery after stroke that may summarise the four functional domains being treated. The secondary outcome measures were chosen according to the ICF because the ICF framework is a comprehensive and person-centred approach that utilizes the concept of human functioning adopted by the World Health Organization. It is based on the idea that health is not just the biological state of the body, but also the ability to take part in daily life and achieve personal goals.

The study aims to demonstrate the non-inferiority (and eventually, the superiority if non-inferiority is established) of rehabilitation treatment integrated with RADTs, compared to only traditional rehabilitation treatment in stroke patients in the recovery of daily life abilities. Secondary objectives include assessing the targeted domains (upper limb, lower limb, balance, cognitive functions) according to the ICF. Moreover, by analysing neurophysiological parameters and factors related to neuroplasticity, along with demographic and clinical data, we can deeply profile patients to identify the factors that predispose them to recovery.

The study also intends to assess the acceptability and usability of robot-assisted rehabilitation for both patients and healthcare providers, develop a predictive model to forecast the effectiveness of robotic treatment and evaluate its economic and organizational sustainability. The ultimate goal is to provide crucial data to guide the large-scale implementation of robot-assisted rehabilitation in stroke patients, considering both clinical effectiveness and economic and organizational aspects. Finally, the data collected in the trial will be made available to the scientific community in accordance with the "Open science" and "FAIR Data".

A limitation of the study is the use of different technologies across various research centres. However, this approach allows us to conduct a trial that more accurately reflects real-world conditions. Given the wide range of existing robotic and technical systems and the constant development of new ones, it is unlikely that all centres offering robotic rehabilitation for stroke patients would use the same technology. Had we standardized the technology across all centres, we would have faced challenges in recruiting a sufficient number of patients, as the number of centres with identical robotic systems within the Fit4MedRob consortium is limited. More importantly, this would have compromised the generalizability of the results. To address the inevitable heterogeneity in the systems used to treat the experimental group, we precisely defined the functional domains that robotic or technological treatments must target, including the lower limb, balance, proximal and distal upper limb segments, and cognitive functions.

Another potential limitation is the inclusion of traditional treatment alongside the specific sessions for each treatment arm (robotic or traditional). In the proposed experiment, patients in the experimental arm will receive both targeted interventions for the five functional domains using one or more robots per domain and traditional treatment. To address this, we ensured that patients in both treatment arms received the same amount of therapy: two daily sessions of traditional rehabilitation in the traditional group, one session of traditional rehabilitation and one session of robotic rehabilitation daily in the robotic group. This design choice stems from our belief that traditional and robotic treatments are complementary rather than mutually exclusive. We are convinced that robotic devices cannot replace the expertise and proven effectiveness of traditional rehabilitation but can serve as valuable tools in the hands of skilled therapists, enabling more precise and personalized treatment.

It is important to note that this trial was designed based on the results of a survey conducted by the Fit4MedRob Consortium, which highlighted an urgent need for large-scale, pragmatic studies focusing on comprehensive recovery of sensorimotor and cognitive functions through a multidomain approach. The survey indicated that many patients require simultaneous rehabilitation across multiple functional domains, necessitating a shift from treating individual domains to adopting a holistic approach, in line with the World Health Organization’s concept of "human functioning." Furthermore, developing sustainable and advanced therapies is crucial to providing effective rehabilitation with robotic systems in a cost-efficient manner for the national health system.

### Ethics and dissemination

This study protocol (version 2, 29/05/2024) was reviewed and approved by the National Ethics Committee for clinical trials of Public Research Bodies (EPR) and other National Public Institutions (CEN) on July 17^th^, 2024 (Prot. PRE BIO CE n. 0027276).

This trial collects personal data, including sensitive categories as per Article 9 of the European regulation on the protection of personal data 2016\679 (General Data Protection Regulation or "GDPR"). The legal basis for data collection and processing is the informed consent of enrolled patients (Article 6 para. 1 letter a, and Article 9 para. 2 letter a GDPR) and the personnel involved in the questionnaires (Article 6 para. 1 letter a GDPR). The latter will only involve personal data as per Article 6 para. 1 of the GDPR, excluding sensitive data.

Upon consent from the subject or their family/caregiver/guardian, data will be pseudonymized and then entered into a computerized database using the REDCap (Research Electronic Data Capture) application installed on an AWS Cloud space managed by the Information Systems of the University of Pavia. During the study, access to the registry will only be granted to the principal investigators of each experimental centre, concerning the data of their Reference centre. Collaborators accessing the computerized database will not be able to trace back the identity of the subjects in any way. Not even the Sponsor will have access to re-identification keys but will have access to all data entered in the REDCap registry (77). Only the principal investigators of each centre will be able to trace back the identities of the enrolled participants at their own centre. The correspondence between the patient’s name and the associated ID will be kept in a protected external file, accessible only to the Principal Investigators of the individual recruitment centres for any clinical follow-up purposes. This file will be deleted once data collection is completed and the quality control of the collected data is carried out. Consequently, at the end of the database establishment, this data will be definitively anonymized by destroying the re-identification keys and potentially removing identifying combinations (for example, patient data linkage with the collection centre) as indicated in recital 26 of the GDPR.

For this reason and in compliance with the principle of minimal data retention, they will be kept for 10 years after the conclusion of the study. Once the aforementioned retention period has expired, the data will be verified for the maintenance of anonymity and possibly kept anonymous so that it continues to be impossible, directly or indirectly, to trace back to the identity of the data subject. These data may thus be reused for subsequent research purposes under Article 89 GDPR and may therefore be retained indefinitely.

Regarding biological samples, for each patient serum samples (aliquots) and whole blood tubes will be identified by reference ID and stored at -20°C until shipment. All samples will be then transferred and analysed at the laboratory located at the Fondazione Don Carlo Gnocchi ONLUS “S. Maria della Provvidenza” Centre, Rome. Samples will be shipped to this laboratory according to specific procedures and maintaining a controlled temperature (dry ice shipment).

The biological material will be utilized exclusively for research purposes related to the execution of the study and it will be analysed only by the personnel of the above-mentioned Centre at Fondazione Don Carlo Gnocchi, Rome. The collected biological material will be stored under secure conditions and according to scientifically recognized procedures. The sample will be preserved with alphanumeric labelling not traceable to the individual patient. Biological data will be processed in accordance with the GDPR and the current national legislation (Legislative Decree 196/2003 and subsequent amendments), as well as the Deontological Rules for statistical or scientific research treatments published pursuant to Article 20, paragraph 4, of Legislative Decree 10 August 2018, No. 101 - December 19, 2018. In particular, the processing, with the aforementioned security measures, will take place in compliance with Article 9 of the Regulation, Article 2-sexies, the prescriptions identified by the Data Protection Authority pursuant to Article 21 of Legislative Decree No. 101 of 2018, and the specific safeguards adopted by the Data Protection Authority data pursuant to Article 2-septies of Legislative Decree 196/2003.

The research team plans to disseminate the trial findings at both national and international conferences, as well as publish them in international journals. Patient data collected during the trial will be anonymized and made available to the scientific community in accordance with principles of "Open Science" and "FAIR Data".

### Patients’ enrolment monitoring

In order to keep the information on patients’ enrolment by every centre up-to-date, a monitoring tool was developed using the REDCap platform. Each centre identified a responsible person who has given credentials for entering data for that centre. Each centre is periodically required to fill in a form reporting the number of patients enrolled so far, possible dropouts, and possible issues. The main objective of this dashboard is to allow the detection of criticalities as soon as possible, in such a way as to take corrective actions on time.

## Data Availability

This is a Clinical research design protocol. No data are reported in the Manuscript.

## Collaborators

The StrokeFit4 study group:

• Laura Cortellini, Stefania Lattanzi, Silvia Scala, Carola Cocco, Francesca Falchini (IRCCS Fondazione Don Carlo Gnocchi);
• Luca Martinis, Ilaria Campese (IRCCS Fondazione Mondino);
• Stefania De Trane, Antonella Laddaga, Matteo Gallotta, Anna Markova (Istituti Clinici Scientifici Maugeri IRCCS);
• Matteo Pittaluga, Filippo Cotellessa, Mehrnaz Hamedani (IRCCS Ospedale Policlinico San Martino).

## Authors’ contributions

*Study conception and design*: Irene Giovanna Aprile, Marco Germanotta, Alessio Fasano, Mariacristina Siotto, Arianna Pavan, Maria Cristina Mauro, Giovanna Nicora, Giuseppina Sgandurra, Alberto Malovini, Maria Letizia Oreni, Nevio Dubbini, Christian Lunetta, Pietro Fiore, Roberto De Icco, Carlo Trompetto, Leopoldo Trieste, Giuseppe Turchetti, Silvana Quaglini. *Wrote the first draft*: Irene Giovanna Aprile, Marco Germanotta, Alessio Fasano. *Critically reviewed the manuscript*: Mariacristina Siotto, Arianna Pavan, Maria Cristina Mauro, Giovanna Nicora, Giuseppina Sgandurra, Alberto Malovini, Maria Letizia Oreni, Nevio Dubbini, Enea Parimbelli, Christian Lunetta, Pietro Fiore, Roberto De Icco, Carlo Trompetto, Leopoldo Trieste, Giuseppe Turchetti, Silvana Quaglini.

## Acknowledgements

We thank Professor John H. Holmes from the University of Pennsylvania Perelman School of Medicine for his help in the initial phase of the trial design.

## Patient and public involvement

This study was designed in response to the needs reported by patients, who have indicated that their expectations have not yet been fully met by existing robotic technologies. These unmet needs were systematically collected through a comprehensive survey conducted by the Fit4MedRob Consortium, which involved both patients and healthcare professionals. The insights gathered from this survey played a pivotal role in shaping the study’s objectives and methodology, ensuring that the research directly addresses the gaps identified by end-users and aligns with the evolving demands of clinical practice.

Moreover, the Italian patients’ association ALICe (Associazione per la Lotta all’Ictus Cerebrale, Association for the Fight Against Brain Stroke) has endorsed the STROKEFIT4 trial. ALICe contributed to the study design and will also play a key role in the dissemination plan.

## References

1. Prange GB, Jannink MJA, Groothuis-Oudshoorn CGM, Hermens HJ, IJzerman MJ. Systematic review of the effect of robot-aided therapy on recovery of the hemiparetic arm after stroke. JRRD. 2006;43(2):171.

2. Riener R. Rehabilitation Robotics. FNT in Robotics. 2012;3(1–2):1–137.

3. Laut J, Porfiri M, Raghavan P. The Present and Future of Robotic Technology in Rehabilitation. Curr Phys Med Rehabil Rep. 2016 Dec;4(4):312–9.

4. Robertson JVG, Roby-Brami A. Augmented feedback, virtual reality and robotics for designing new rehabilitation methods. In: Didier JP, Bigand E, editors. Rethinking physical and rehabilitation medicine: New technologies induce new learning strategies [Internet]. Paris: Springer; 2010 [cited 2024 Apr 3]. p. 223–45. Available from: 10.1007/978-2-8178-0034-9_12

5. Germanotta M, Cruciani A, Pecchioli C, Loreti S, Spedicato A, Meotti M, et al. Reliability, validity and discriminant ability of the instrumental indices provided by a novel planar robotic device for upper limb rehabilitation. J NeuroEngineering Rehabil. 2018 Dec;15(1):39.

6. Germanotta M, Gower V, Papadopoulou D, Cruciani A, Pecchioli C, Mosca R, et al. Reliability, validity and discriminant ability of a robotic device for finger training in patients with subacute stroke. J NeuroEngineering Rehabil. 2020 Dec;17(1):1.

7. Camardella C, Cappiello G, Curto Z, Germanotta M, Aprile I, Mazzoleni S, et al. A Random Tree Forest decision support system to personalize upper extremity robot-assisted rehabilitation in stroke: a pilot study. IEEE Int Conf Rehabil Robot. 2022 Jul;2022:1–6.

8. Pavan A, Fasano A, Cortellini L, Lattanzi S, Papadopoulou D, Insalaco S, et al. Implementation of a robot-mediated upper limb rehabilitation protocol for a customized treatment after stroke: A retrospective analysis. NeuroRehabilitation. 2024 Mar 5;

9. Aprile I, Germanotta M, Cruciani A, Loreti S, Pecchioli C, Cecchi F, et al. Upper Limb Robotic Rehabilitation After Stroke: A Multi centre, Randomized Clinical Trial. J Neurol Phys Ther. 2020 Jan;44(1):3–14.

10. Rodgers H, Bosomworth H, Krebs HI, Van Wijck F, Howel D, Wilson N, et al. Robot assisted training for the upper limb after stroke (RATULS): a multicentre randomised controlled trial. The Lancet. 2019 Jul;394(10192):51–62.

11. Wu J, Cheng H, Zhang J, Yang S, Cai S. Robot-Assisted Therapy for Upper Extremity Motor Impairment After Stroke: A Systematic Review and Meta-Analysis. Physical Therapy. 2021 Apr 4;101(4):pzab010.

12. Yang X, Shi X, Xue X, Deng Z. Efficacy of Robot-Assisted Training on Rehabilitation of Upper Limb Function in Patients With Stroke: A Systematic Review and Meta-analysis. Archives of Physical Medicine and Rehabilitation. 2023 Sep;104(9):1498–513.

13. Mehrholz J, Pohl M, Platz T, Kugler J, Elsner B. Electromechanical and robot-assisted arm training for improving activities of daily living, arm function, and arm muscle strength after stroke. Cochrane Stroke Group, editor. Cochrane Database of Systematic Reviews [Internet]. 2018 Sep 3 [cited 2024 Apr 3];2018(9). Available from: https://doi.wiley.com/10.1002/14651858.CD006876.pub5

14. Aprile I, Guardati G, Cipollini V, Papadopoulou D, Mastrorosa A, Castelli L, et al. Robotic Rehabilitation: An Opportunity to Improve Cognitive Functions in Subjects With Stroke. An Explorative Study. Front Neurol. 2020;11:588285.

15. Negrini S, Selb M, Kiekens C, Todhunter-Brown A, Arienti C, Stucki G, et al. Rehabilitation definition for research purposes. A global stakeholders’ initiative by Cochrane Rehabilitation. Eur J Phys Rehabil Med. 2022 Jun;58(3):333–41.

16. Santoro M, Siotto M, Germanotta M, Bray E, Mastrorosa A, Galli C, et al. Bdnf rs6265 polymorphism and its methylation in patients with stroke undergoing rehabilitation. Int J Mol Sci. 2020;21(22):1–11.

17. Mojtabavi H, Shaka Z, Momtazmanesh S, Ajdari A, Rezaei N. Circulating brain-derived neurotrophic factor as a potential biomarker in stroke: a systematic review and meta-analysis. J Transl Med. 2022 Mar 14;20(1):126.

18. Pekny M, Wilhelmsson U, Stokowska A, Tatlisumak T, Jood K, Pekna M. Neurofilament Light Chain (NfL) in Blood—A Biomarker Predicting Unfavourable Outcome in the Acute Phase and Improvement in the Late Phase after Stroke. Cells. 2021 Jun 18;10(6):1537.

19. Liu Z, Li Y, Cui Y, Roberts C, Lu M, Wilhelmsson U, et al. Beneficial effects of gfap/vimentin reactive astrocytes for axonal remodeling and motor behavioral recovery in mice after stroke. Glia. 2014 Dec;62(12):2022–33.

20. Huppertz V, Guida S, Holdoway A, Strilciuc S, Baijens L, Schols JMGA, et al. Impaired Nutritional Condition After Stroke From the Hyperacute to the Chronic Phase: A Systematic Review and Meta-Analysis. Front Neurol. 2022 Feb 1;12:780080.

21. Turchetti G, Spadoni E, Geisler EE. Health technology assessment. Evaluation of biomedical innovative technologies. IEEE Eng Med Biol Mag. 2010;29(3):70–6.

22. Flynn N, Froude E, Cooke D, Dennis J, Kuys S. The sustainability of upper limb robotic therapy for stroke survivors in an inpatient rehabilitation setting. Disability and Rehabilitation. 2022 Nov 20;44(24):7522– 7.

23. Turchetti G, Vitiello N, Trieste L, Romiti S, Geisler E, Micera S. Why effectiveness of robot-mediated neurorehabilitation does not necessarily influence its adoption. IEEE Rev Biomed Eng. 2014;7:143–53.

24. Bertolini A, Salvini P, Pagliai T, Morachioli A, Acerbi G, Trieste L, et al. On Robots and Insurance. Int J of Soc Robotics. 2016 Jun 1;8(3):381–91.

25. Ford I, Norrie J. Pragmatic Trials. Drazen JM, Harrington DP, McMurray JJV, Ware JH, Woodcock J, editors. N Engl J Med. 2016 Aug 4;375(5):454–63.

26. World Health Organization. IFC: International Classification of Functioning, Disability and Health. 2001.

27. Chan AW, Tetzlaff JM, Altman DG, Laupacis A, Gøtzsche PC, Krleža-Jerić K, et al. SPIRIT 2013 Statement: Defining Standard Protocol Items for Clinical Trials. Ann Intern Med. 2013 Feb 5;158(3):200– 7.

28. Bernhardt J, Hayward KS, Kwakkel G, Ward NS, Wolf SL, Borschmann K, et al. Agreed Definitions and a Shared Vision for New Standards in Stroke Recovery Research: The Stroke Recovery and Rehabilitation Roundtable Taskforce. Neurorehabil Neural Repair. 2017 Sep;31(9):793–9.

29. Cecchi F, Carrabba C, Bertolucci F, Castagnoli C, Falsini C, Gnetti B, et al. Transcultural translation and validation of Fugl–Meyer assessment to Italian. Disability and Rehabilitation. 2021 Dec 4;43(25):3717– 22.

30. Holden MK, Gill KM, Magliozzi MR, Nathan J, Piehl-Baker L. Clinical Gait Assessment in the Neurologically Impaired. Physical Therapy. 1984 Jan 1;64(1):35–40.

31. Berg K, Wood-Dauphinee S, Williams JI. The Balance Scale: reliability assessment with elderly residents and patients with an acute stroke. Scand J Rehabil Med. 1995 Mar;27(1):27–36.

32. Nasreddine ZS, Phillips NA, Bédirian V, Charbonneau S, Whitehead V, Collin I, et al. The Montreal Cognitive Assessment, MoCA: A Brief Screening Tool For Mild Cognitive Impairment. J American Geriatrics Society. 2005 Apr;53(4):695–9.

33. Bohannon RW, Smith MB. Interrater Reliability of a Modified Ashworth Scale of Muscle Spasticity. Physical Therapy. 1987 Feb 1;67(2):206–7.

34. Banks JL, Marotta CA. Outcomes validity and reliability of the modified Rankin scale: implications for stroke clinical trials: a literature review and synthesis. Stroke. 2007 Mar;38(3):1091–6.

35. Nucci M, Mapelli D, Mondini S. Cognitive Reserve Index questionnaire (CRIq): a new instrument for measuring cognitive reserve. Aging Clin Exp Res. 2012 Jun;24(3):218–26.

36. Friedenreich CM, Courneya KS, Bryant HE. The Lifetime Total Physical Activity Questionnaire: development and reliability: Medicine &amp Science in Sports &amp Exercise. 1998 Feb;30(2):266–74.

37. Brott T, Adams HP, Olinger CP, Marler JR, Barsan WG, Biller J, et al. Measurements of acute cerebral infarction: a clinical examination scale. Stroke. 1989 Jul;20(7):864–70.

38. Suda M, Kawakami M, Okuyama K, Ishii R, Oshima O, Hijikata N, et al. Validity and Reliability of the Semmes-Weinstein Monofilament Test and the Thumb Localizing Test in Patients With Stroke. Frontiers in Neurology [Internet]. 2021 [cited 2023 Oct 20];11. Available from: https://www.frontiersin.org/articles/10.3389/fneur.2020.625917

39. Mead G, Lynch J, Greig C, Young A, Lewis S, Sharpe M. Evaluation of Fatigue Scales in Stroke Patients. Stroke. 2007 Jul;38(7):2090–5.

40. Salvi F, Miller MD, Grilli A, Giorgi R, Towers AL, Morichi V, et al. A Manual of Guidelines to Score the Modified Cumulative Illness Rating Scale and Its Validation in Acute Hospitalized Elderly Patients. J American Geriatrics Society. 2008 Oct;56(10):1926–31.

41. Blaylock A, Cason CL. Discharge planning predicting patients’ needs. J Gerontol Nurs. 1992 Jul;18(7):5– 9.

42. McGarvey SR, Morrey BF, Askew LJ, An KN. Reliability of isometric strength testing. Temporal factors and strength variation. Clin Orthop Relat Res. 1984 May;(185):301–5.

43. Rubenstein LZ, Harker JO, Salva A, Guigoz Y, Vellas B. Screening for Undernutrition in Geriatric Practice: Developing the Short-Form Mini-Nutritional Assessment (MNA-SF). The Journals of Gerontology Series A: Biological Sciences and Medical Sciences. 2001 Jun 1;56(6):M366–72.

44. Crary MA, Mann GDC, Groher ME. Initial Psychometric Assessment of a Functional Oral Intake Scale for Dysphagia in Stroke Patients. Archives of Physical Medicine and Rehabilitation. 2005 Aug;86(8):1516–20.

45. Malmstrom TK, Morley JE. SARC-F: a simple questionnaire to rapidly diagnose sarcopenia. Journal of the American Medical Directors Association. 2013;14(8):531–2.

46. Beck AT, Steer RA, Brown G. Beck depression inventory–II. Psychological assessment. 1996;

47. Zigmond AS, Snaith RP. The Hospital Anxiety and Depression Scale. Acta Psychiatr Scand. 1983 Jun;67(6):361–70.

48. McAuley E, Duncan T, Tammen VV. Psychometric properties of the Intrinsic Motivation Inventory in a competitive sport setting: a confirmatory factor analysis. Res Q Exerc Sport. 1989 Mar;60(1):48–58.

49. Winstein CJ, Stein J, Arena R, Bates B, Cherney LR, Cramer SC, et al. Guidelines for Adult Stroke Rehabilitation and Recovery: A Guideline for Healthcare Professionals From the American Heart Association/American Stroke Association. Stroke [Internet]. 2016 Jun [cited 2024 Apr 5];47(6). Available from: https://www.ahajournals.org/doi/10.1161/STR.0000000000000098

50. Hunter SM, Crome P, Sim J, Donaldson C, Pomeroy VM. Development of treatment schedules for research: a structured review to identify methodologies used and a worked example of ‘mobilisation and tactile stimulation’ for stroke patients. Physiotherapy. 2006 Dec;92(4):195–207.

51. Donaldson C, Tallis RC, Pomeroy VM. A treatment schedule of conventional physical therapy provided to enhance upper limb sensorimotor recovery after stroke: Expert criterion validity and intra-rater reliability. Physiotherapy. 2009 Jun;95(2):110–9.

52. Linee Guida SPREAD VII edizione. In: isa-aii [Internet]. 2012 [cited 2024 Apr 5]. Available from: https://isa-aii.com/linee-guida-spread-vii-edizione/

53. Overview | Stroke rehabilitation in adults | Guidance | NICE [Internet]. NICE; 2023 [cited 2024 Apr 5]. Available from: https://www.nice.org.uk/guidance/NG236

54. Norrving B, Barrick J, Davalos A, Dichgans M, Cordonnier C, Guekht A, et al. Action Plan for Stroke in Europe 2018-2030. Eur Stroke J. 2018 Dec;3(4):309–36.

55. Hoffmann TC, Glasziou PP, Boutron I, Milne R, Perera R, Moher D, et al. Better reporting of interventions: template for intervention description and replication (TIDieR) checklist and guide. BMJ. 2014 Mar 7;348:g1687.

56. Smith A. Symbol Digit Modalities Test. Western psychological services Los Angeles [Internet]. 1973 [cited 2024 Apr 4]; Available from: https://doi.apa.org/doi/10.1037/t27513-000

57. Demeurisse G, Demol O, Robaye E. Motor evaluation in vascular hemiplegia. Eur Neurol. 1980;19(6):382–9.

58. Downie WW, Leatham PA, Rhind VM, Wright V, Branco JA, Anderson JA. Studies with pain rating scales. Annals of the Rheumatic Diseases. 1978 Aug 1;37(4):378–81.

59. Bouhassira D, Attal N, Alchaar H, Boureau F, Brochet B, Bruxelle J, et al. Comparison of pain syndromes associated with nervous or somatic lesions and development of a new neuropathic pain diagnostic questionnaire (DN4). Pain. 2005 Mar;114(1):29–36.

60. Lyle RC. A performance test for assessment of upper limb function in physical rehabilitation treatment and research: International Journal of Rehabilitation Research. 1981 Dec;4(4):483–92.

61. Steele B. Timed Walking Tests of Exercise Capacity in Chronic Cardiopulmonary Illness: Journal of Cardiopulmonary Rehabilitation. 1996 Jan;16(1):25–33.

62. Feng YS, Kohlmann T, Janssen MF, Buchholz I. Psychometric properties of the EQ-5D-5L: a systematic review of the literature. Qual Life Res. 2021 Mar;30(3):647–73.

63. Van Putten MJAM. The revised brain symmetry index. Clinical Neurophysiology. 2007 Nov;118(11):2362–7.

64. Sheorajpanday RVA, Nagels G, Weeren AJTM, Van Putten MJAM, De Deyn PP. Reproducibility and clinical relevance of quantitative EEG parameters in cerebral ischemia: A basic approach. Clinical Neurophysiology. 2009 May;120(5):845–55.

65. Claassen J, Hirsch LJ, Kreiter KT, Du EY, Sander Connolly E, Emerson RG, et al. Quantitative continuous EEG for detecting delayed cerebral ischemia in patients with poor-grade subarachnoid hemorrhage. Clinical Neurophysiology. 2004 Dec;115(12):2699–710.

66. Mathiowetz V, Volland G, Kashman N, Weber K. Adult norms for the Box and Block Test of manual dexterity. Am J Occup Ther. 1985 Jun;39(6):386–91.

67. Watson MJ. Refining the Ten-metre Walking Test for Use with Neurologically Impaired People. Physiotherapy. 2002 Jul;88(7):386–97.

68. Finch AP, Meregaglia M, Ciani O, Roudijk B, Jommi C. An EQ-5D-5L value set for Italy using videoconferencing interviews and feasibility of a new mode of administration. Social Science & Medicine. 2022 Jan;292:114519.

69. Venkatesh V, Bala H. Technology Acceptance Model 3 and a Research Agenda on Interventions. Decision Sciences. 2008 May;39(2):273–315.

70. Van Laar D, Edwards JA, Easton S. The Work-Related Quality of Life scale for healthcare workers. Journal of Advanced Nursing. 2007 Nov;60(3):325–33.

71. Shima N, Miyamoto K, Shibata M, Nakashima T, Kaneko M, Shibata N, et al. Activities of daily living status and psychiatric symptoms after discharge from an intensive care unit: a single-centre 12-month longitudinal prospective study. Acute Medicine & Surgery. 2020 Jan;7(1):e557.

72. Andrew MK, MacDonald S, Godin J, McElhaney JE, LeBlanc J, Hatchette TF, et al. Persistent Functional Decline Following Hospitalization with Influenza or Acute Respiratory Illness. J American Geriatrics Society. 2021 Mar;69(3):696–703.

73. Hsieh YW, Wang CH, Wu SC, Chen PC, Sheu CF, Hsieh CL. Establishing the Minimal Clinically Important Difference of the Barthel Index in Stroke Patients. Neurorehabil Neural Repair. 2007 May;21(3):233–8.

74. Faul F, Erdfelder E, Lang AG, Buchner A. G*Power 3: A flexible statistical power analysis program for the social, behavioral, and biomedical sciences. Behavior Research Methods. 2007 May;39(2):175–91.

75. White IR, Horton NJ, Carpenter J, Statistics RIMAS, Pocock SJ. Strategy for intention to treat analysis in randomised trials with missing outcome data. BMJ. 2011 Feb 7;342(feb07 1):d40–d40.

76. White IR, Carpenter J, Horton NJ. Including all individuals is not enough: Lessons for intention-to-treat analysis. Clinical Trials. 2012 Aug;9(4):396–407.

77. Harris PA, Taylor R, Minor BL, Elliott V, Fernandez M, O’Neal L, et al. The REDCap consortium: Building an international community of software platform partners. Journal of Biomedical Informatics. 2019 Jul;95:103208.

